# The gray swan: model-based assessment of the risk of sudden failure of hybrid immunity to SARS-CoV-2

**DOI:** 10.1101/2023.02.26.23286471

**Authors:** Madison Stoddard, Lin Yuan, Sharanya Sarkar, Debra Van Egeren, Laura F. White, Arijit Chakravarty

**Affiliations:** Fractal Therapeutics, Cambridge, MA, USA; Dartmouth College, Hanover, NH, USA; Stanford University School of Medicine, Stanford, CA, USA; Boston University, Boston, MA, USA

## Abstract

In the fourth year of the COVID-19 pandemic, public health authorities worldwide have adopted a strategy of learning to live with SARS-CoV-2. This has involved the removal of measures for limiting viral spread, resulting in a large burden of recurrent SARS-CoV-2 infections. Crucial for managing this burden is the concept of the so-called wall of hybrid immunity, through repeated reinfections and vaccine boosters, to reduce the risk of severe disease and death. Protection against both infection and severe disease is provided by the induction of neutralizing antibodies (nAbs) against SARS-CoV-2. However, pharmacokinetic (PK) waning and rapid viral evolution both degrade nAb binding titers. The recent emergence of variants with strongly immune evasive potential against both the vaccinal and natural immune responses raises the question of whether the wall of population-level immunity can be maintained in the face of large jumps in nAb binding potency. Here we use an agent-based simulation to address this question. Our findings suggest large jumps in viral evolution may cause failure of population immunity resulting in sudden increases in mortality. As a rise in mortality will only become apparent in the weeks following a wave of disease, reactive public health strategies will not be able to provide meaningful risk mitigation. Learning to live with the virus could thus lead to large death tolls with very little warning. Our work points to the importance of proactive management strategies for the ongoing pandemic, and to the need for multifactorial approaches to COVID-19 disease control.

## Introduction

Over the course of the ongoing COVID-19 pandemic, the strategy for the management of the morbidity and mortality burden of SARS-CoV-2 has shifted significantly. The initial perception was that vaccines could be employed to achieve herd immunity to SARS-CoV-2 ^1–5^, based on clinical trial data from vaccine manufacturers demonstrating strong vaccinal protection against symptomatic disease ^6–8^. Real-world studies failed to confirm this effect, and rapid declines in vaccinal efficacy against infection (VE_i_) ^9,10^ driven by waning antibody titers ^11–14^ and viral immune evasion ^14–18^, undermined the promise of herd immunity as a solution to the COVID-19 pandemic. With herd immunity off the table, the strategy pivoted to relying on vaccinal efficacy against severe disease (VE_s_) to keep the death toll down ^19–22^. While VE_s_ was very high in the early stages of vaccine deployment, ^6–8^, continued viral evolution has substantially degraded VE_s_, ^23–26^, undermining this strategy as well (See ^27^ Supplementary Section S1B for more details).

Thus, the public health strategy for COVID-19 now focuses on relying on repeated infections and vaccinations as a means of keeping the death toll down in the teeth of widespread viral transmission (“hybrid immunity”) ^28^. This approach relies on observations indicating that the combination of natural infection and vaccination provides protection against mild symptomatic disease that is similar, or modestly better than that by infection-induced or vaccine-induced immunity alone ^29–34^. In response to the promotion of hybrid immunity by public health authorities ^35^ (see slides 13-18, 15,22), ^36–39^, and media ^40–43^, vaccinal uptake has also declined sharply, with only 18% of adults in the United States, for example, having received the updated booster (January 2023) ^44^.

Unfortunately, the concept of hybrid immunity has been used by some to argue for the removal of nonpharmaceutical interventions (NPIs) ^41,45–47^, inevitably leading to more infections. There are few NPIs mandated in any setting at this point ^48–50^, making it very challenging for individuals to avoid regular infection by SARS-CoV-2 ^51^. At the same time repeat reinfections are expected to lead to a heavy burden of acute and postacute (“long covid”) disease as a result of COVID-19 infections ^51^. While increasing reliance on natural immunity is a trivial consequence of sharply declining vaccinal uptake, it is unclear whether this will offer protection against new variants. For example, recent studies show that the immune response from an Omicron BA.1 infection is almost completely evaded by Omicron BA.5 ^52,53^, while the immune response generated by Omicron BA.5 evaded almost completely by the XBB.1.5 subvariant ^54^.

The rapid evolution of the viral spike protein to evade neutralizing antibodies (nAbs) (referred to here as “immune evasion”) during the current pandemic was predicted by us and others in the fall of 2020 ^55–57^. Positive and negative selection, as well as convergent evolution, have been observed in the viral spike ^58–60^, confirming the clinical significance of the nAb response. Variants of concern in particular have evolved via a process that implicates an acceleration of the substitution rate ^61^, at least in part because of prolonged infections leading to intrahost evolution ^62,63^. The deployment of vaccines in the winter of 2020 led to an acceleration of the evolutionary rate ^64^, and vaccines are not predicted to be able to slow the rate of this evolution going forward ^65^. Using epidemiologic terminology, evolutionary changes in the viral spike protein due to natural selection can be described in terms of the scale of the functional change. Small changes (with a small number of amino acids resulting in minor functional differences) are referred to as “antigenic drift”. On the other hand, large changes in amino acid sequence resulting in abrupt, major functional differences are referred to as “antigenic shift” ^66^.

In fact, as NPIs have been abandoned worldwide, and case counts have risen, antigenic drift has accelerated ^67^, consistent with the early predictions ^55,57^. Notably, the emergence of the Omicron BA.1 variant in the winter of 2021 led to an antigenic shift (a large jump, both in terms of phylogenetic sequence (^68^, Fig.1) and antigenic immune evasion (^68^, Fig. S4) relative to the Delta variant and others that preceded it). An even larger jump (in both sequence and antigenic distance) has occurred recently with the emergence of the XBB/XBB.1 subvariants (^68^, Fig. 1 and Fig. S4). (Interestingly, the XBB/XBB.1 lineage is now antigenically more distant from WT SARS-CoV-2 than WT SARS-CoV-2 is from SARS-CoV-1 ^68^). Consistent with this extreme antigenic distance, nAb titers against XBB subvariants are lower by 66- to 155-fold than against WT ^68^. Despite this strong ability to evade the nAb response, the XBB/XBB.1 lineage has a growth advantage compared to earlier variants and its tight binding affinity for ACE2 will both facilitate its transmissibility and provide an evolutionary buffer enabling further immune evasive mutations ^54^.

**Figure 1:**
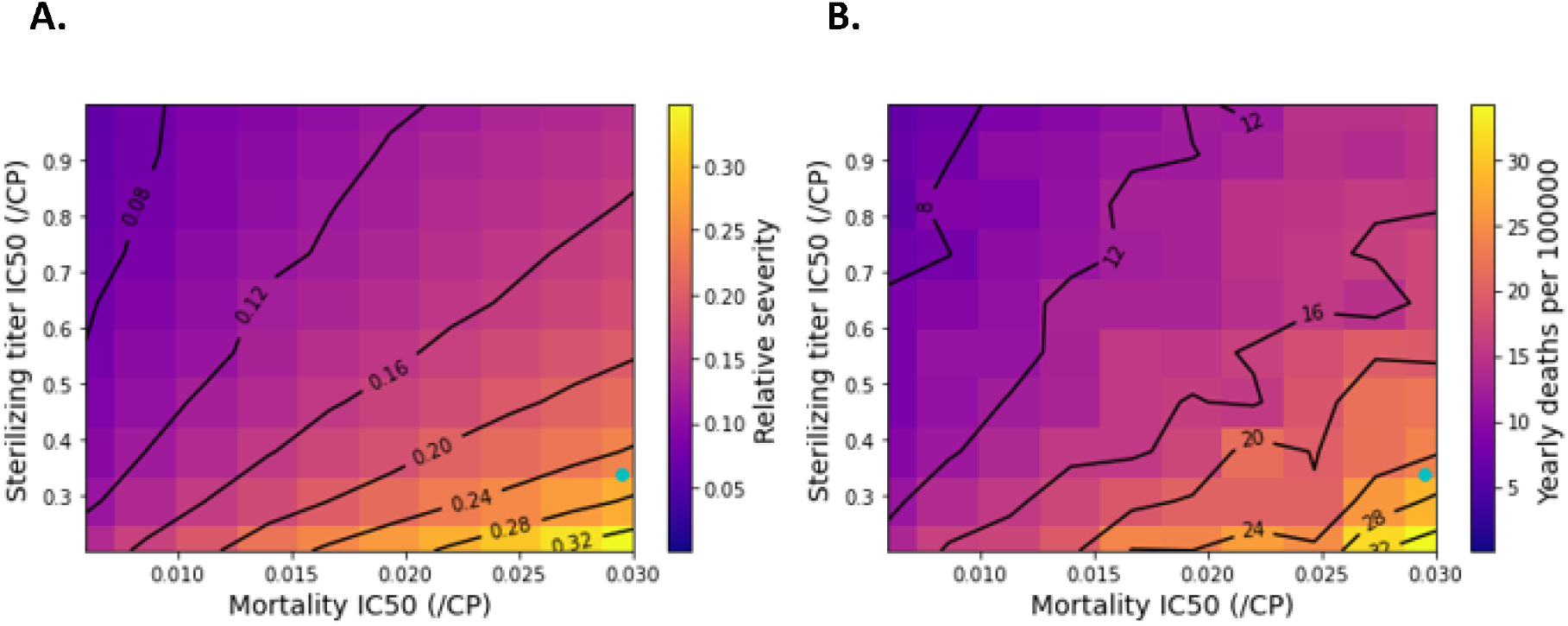
In the absence of immune evasion, immunity could moderate COVID-19 death tolls. Sensitivity sweep for the **A)** fractional severity of COVID-19 reinfections relative to naïve infections and **B)** model-predicted yearly COVID-19 deaths per 100,000. IC_50_s for protection from infection (sterilizing immunity) and death (mortality) are swept over their uncertainty ranges and expressed relative to peak convalescent titer (/CP). Blue point represents best-estimate parameter values.

At this point, the correlates of immune protection against SARS-CoV-2 are well understood. Neutralizing antibody (nAb) titers have been validated as a correlate of immune protection ^10,69–71^, for SARS-CoV-2, both for natural and vaccinal immunity. nAb titers have been demonstrated to predict waning immunity (both natural and vaccinal) driven by pharmacokinetics as well as evolution (See ^27^, Supplementary Section S1H for details). On the other hand, a robust body of evidence argues against a role for T cells in providing protection against severe disease, despite the observed durability of T-cell responses ^72–74^, (See ^27^, Supplementary Section S1 for further discussion on the role of T cells in vaccinal and natural immunity to SARS-CoV-2).

The recent emergence of variants with strongly immune evasive potential against both the vaccinal and natural immune responses raises the question of whether the “wall of immunity” will be maintained in the face of antigenic shifts capable of causing large drops in nAb binding potency ^75^. To address this question, we developed an agent-based modeling framework based on an SEIRS (susceptible-exposed-infected-recovered-susceptible) epidemiological framework, driven by a longitudinal PK/PD model of nAb kinetics. Our model combines population heterogeneity in the durability of the nAb response with the dose-response relationships linking nAb titers to protection from severe disease. We have used this modeling framework to examine the impact of viral immune evasion on mortality outcomes. In particular, we examined both the steady attrition of nAb binding potency due to antigenic drift (arising from within-clade viral evolution) and sudden antigenic shifts due to the emergence of new viral clades, analogous to the Omicron BA.1 and XBB.1.5 shifts). We further examine the impact of altered vaccination schedules on these outcomes.

## Results

### In the absence of viral immune evasion, immunity provides protection against severe COVID

To evaluate the hypothesis that the build-up of immunity under endemic SARS-CoV-2 transmission may reduce the severity of COVID-19, we simulated steady-state COVID-19 outcomes in a population reflecting the United States. In the first set of simulations, we assumed there is no immune evasion due to viral mutation, and thus all nAb decay is due to waning immunity in individuals (Figure 1). Due to uncertainty in the IC_50_s for nAb protection from infection and mortality, we swept these parameters over their plausible ranges (see Methods section for discussion of sweep ranges). This results in a range of possible scenarios. Under all simulated conditions, reinfections are expected to be significantly less severe than naïve infections (Figure 1A). Under our best-estimate parameters, reinfection severity is approximately 0.25 (25% of naïve severity). However, if the IC_50_ for protection from infection is higher and/or the IC_50_ for protection from mortality is lower, reinfection severity could be as low as 0.06, suggesting that hybrid immunity would be expected to provide very robust protection against severe disease in the absence of viral immune evasion.

While reinfection severity depends on the two IC_50_ parameters above, the model-predicted number of yearly infections does not (Figure S1). Approximately 46,000 yearly SARS-CoV-2 infections per 100,000 population are predicted. For the US population of 330 million ^76^, this is equivalent to approximately 150 million SARS-CoV-2 infections annually. The death toll associated with these infections depends on the reinfection severity. Under best-estimate conditions, 25 yearly deaths per 100,000 are predicted (Figure 1B). This corresponds to 82,500 US COVID-19 deaths annually.

### Steady viral immune evasion under antigenic drift leads to more infections, increasing total mortality

Of course, SARS-CoV-2 immune evasion is proceeding at a rapid pace ^64,67^ that is not expected to decrease under conditions of widespread transmission ^65^. To examine the impact of antigenic drift on the robustness of hybrid immunity, we reimplemented the analyses above under the assumption of a continuous rate of immune evasion that degrades nAb potency with a half-life of 73 days (Figure 2) ^51,77^. Intriguingly, the impact of this antigenic drift on reinfection severity is small (Figure 2A). However, the impact on infection counts is vast: roughly 163,000 yearly infections are predicted per 100,000 people (indicating that many people are infected more than once yearly, Figure S2). Immune evasion under conditions of antigenic drift more than triples the expected number of infections. This increase in infections translates to increased death tolls: 70 COVID-19 deaths per 100,000 yearly, or 231,000 annual US deaths. Although vaccinal and natural immunity may reduce the severity of individual infections, the vast transmission of SARS-CoV-2 is expected to continue to drive very significant death tolls.

**Figure 2:**
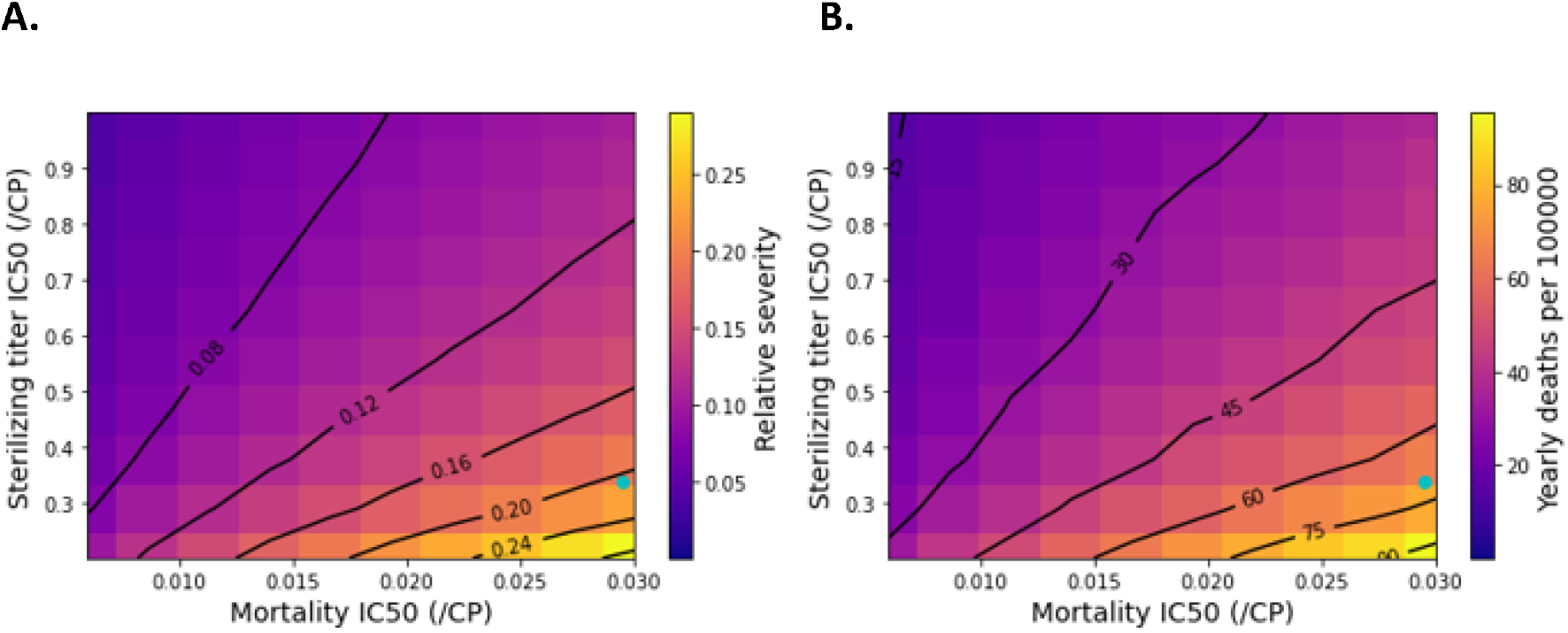
Steady immune evasion substantially increases COVID-19 death tolls with only a minor impact on reinfection severity. Sensitivity sweep for the **A)** fractional severity of COVID-19 reinfections relative to naïve infections and **B)** model-predicted yearly COVID-19 deaths per 100,000. IC_50_s for protection from infection (sterilizing immunity) and death (mortality) are swept over their uncertainty ranges. Blue point represents best-estimate parameter values.

The relationship between the rate of immune evasion and COVID-19 outcomes is further elucidated in Figure 3. Figure 3A demonstrates that the rate of continuous immune evasion has little impact on reinfection severity. However, as the rate of immune evasion increases (and the half-life of immune evasion decreases), SARS-CoV-2 infections (Figure 3B) and deaths (Figure 3C) increase steeply.

**Figure 3:**
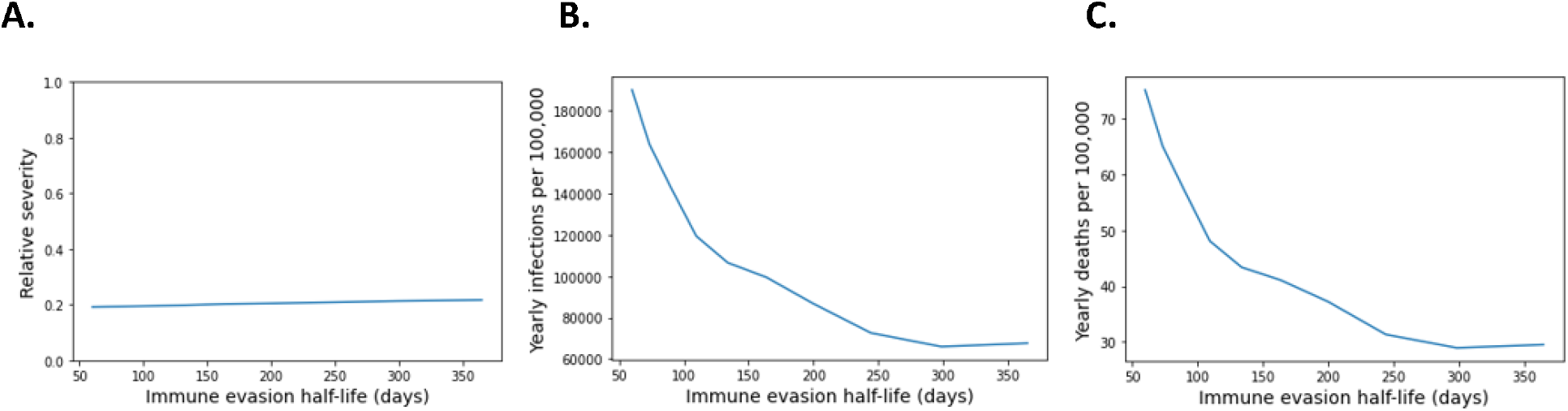
Higher rates of continuous immune evasion drive increased yearly SARS-CoV-2 infections and deaths. **A**. Relative severity of reinfections as a function of immune evasion half-life. Relative severity is minimally impacted by immune-evasion half-life. See Methods for the definition of relative severity. **B**. Steady-state yearly SARS-CoV-2 infections as a function of immune evasion half-life. **C**. Steady-state yearly COVID-19 deaths as a function of immune evasion half-life.

### Large drops in nAb potency due to antigenic shift can transiently increase transmission and apparent IFR

Phylogenetic evidence suggests that SARS-CoV-2’s overall rate of evolution is driven in large part by the emergence of variants or subvariants with many simultaneous amino acid changes^63,78–80^. While a continuous rate of immune evasion is likely sufficient to model behavior under conditions of antigenic drift via between-host evolution, variant emergence due to antigenic shift results in sudden, virtually discrete changes in nAb potency. To determine COVID-19 outcomes under conditions of variant emergence, we evaluated the impact of sudden shifts in nAb potency driven by viral evolution accompanied by possible changes in viral R_0_.

In Figure 4A, we explore the impact of antigenic shift on the relative severity of SARS-CoV-2 reinfections using best-estimate parameters. Although immunological protection is not fully eroded even with a 100-fold drop in nAb potency, immune evasion can significantly increase the apparent fatality rate of reinfections. For example, infections in the first wave of transmission of a variant with R_0_ comparable to omicron BA.1 (R_0_ = 8, ^81,82^) and a 50-fold reduction in potency of pre-existing nAbs would have an increased relative severity of 50%. This represents a 2.5-fold increase in apparent IFR relative to baseline, endemic conditions (Figure 4B). On the other hand, variants exerting smaller drops in nAb potency (e.g. less than 10-fold) are expected to have a less pronounced impact on IFR (Figures S3A and S3B). Such immune evading variants have the potential to spread rapidly after their emergence, with R_t_ estimates exceeding 5 in some cases (Figures 4C and S3C).

**Figure 4:**
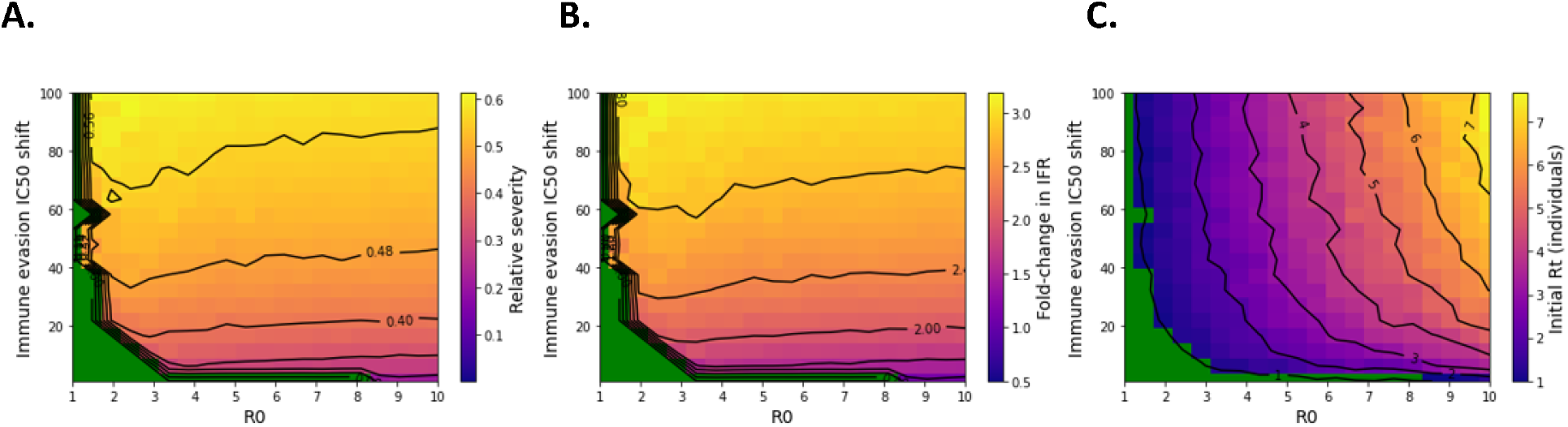
Emergence of an immune-evading SARS-CoV-2 variant can increase IFR and R_t_ compared to steady-state conditions. In these simulations, we used best-estimate values for endemic model parameters while varying the R_0_ and immune evasion IC_50_ shift of the invading variant. Green region represents extinction of the invading variant under conditions of pre-existing immunity. **A**. Severity of immune evasive variant infections compared to naïve during the first wave of spread. **B**. Transient fold-change in IFR during immune-evasive variant wave. C R_t_ of the novel variant at the time of introduction.

Although strongly immune evading variants are likely to spread and impact IFR, vaccines still improve population-level outcomes. As shown in Figure S4, disease severity would increase in the absence of vaccines. Increasing vaccine compliance and booster frequency would not prevent transmission of the most immune evasive variants but could blunt their impact on mortality (Figure S5).

### Rapid variant outbreak kinetics are accompanied by increases in apparent IFR

In Figure 5, we explored the relationship between relative severity and R_t_ of novel variants during their initial spread. Although moderately transmissible novel variants have a range of impacts on IFR, the most transmissible variants all increase IFR by 2 to 3-fold. This suggests that while highly rapidly spreading novel variants are very likely to increase reinfection severity, this outcome is also possible with immune evasive variants that transmit less efficiently upon emergence. The immune evasion potential of these variants determines their impact on severe disease. However, the existence of highly-transmissible strains with high reinfection severity implies that antigenic shifts represent a pernicious public health threat at this stage of the pandemic.

**Figure 5:**
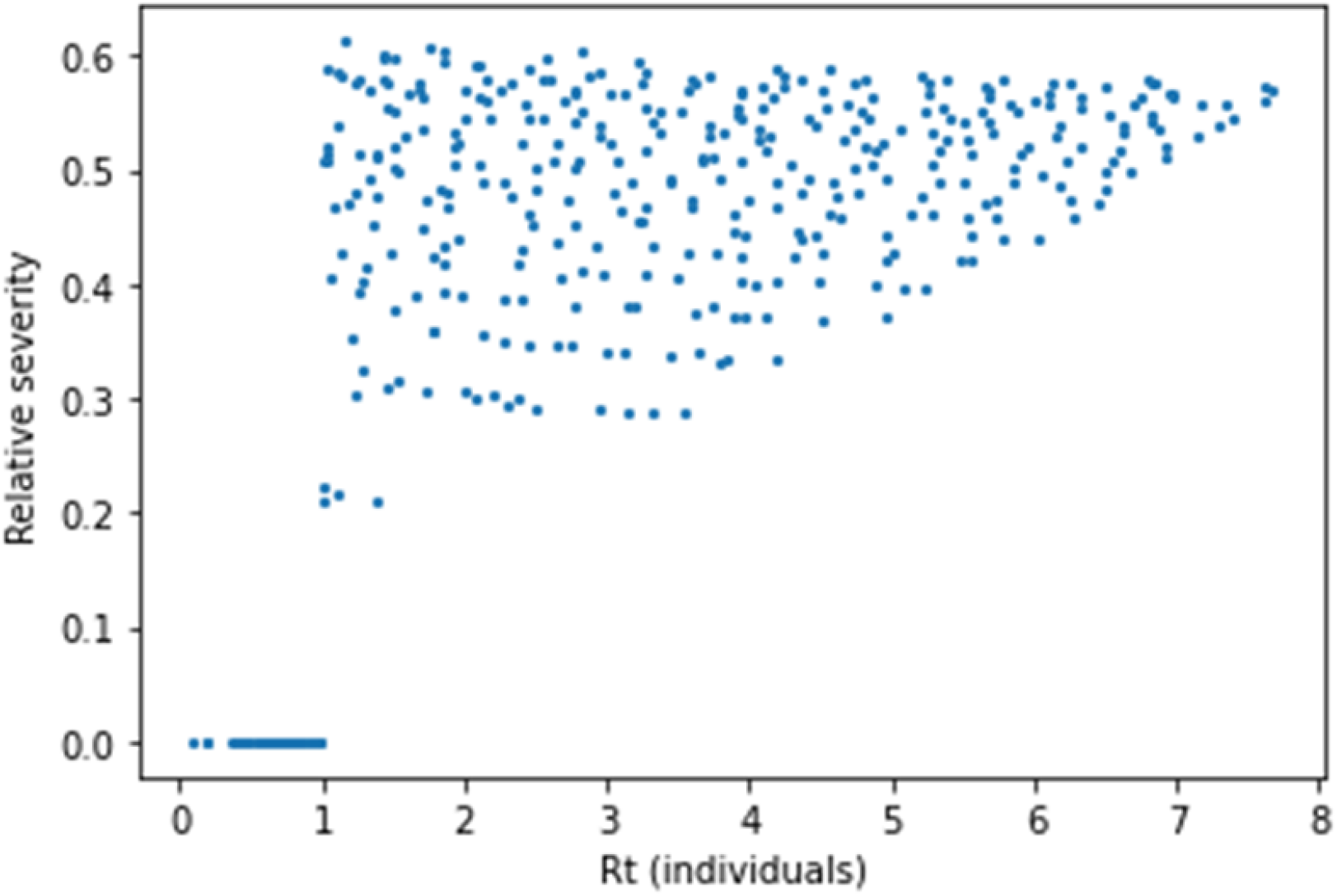
Variants with greater effective transmissibility (R_t_) upon introduction increase IFR to a greater degree. Although moderately transmissible novel variants have a range of impacts on IFR, the most transmissible variants all increase IFR by 2 to 3-fold.

### Greater population-level immunological protection from severe disease is accompanied by greater vulnerability to changes in IFR

To test the impact of our assumptions regarding the relationships between nAb titer and protection against SARS-CoV-2 infection and mortality, we reimplemented the analyses of Figure 4 while assuming a higher IC_50_ for protection from infection (81% of peak convalescent titer, CP) and a lower IC_50_ for protection from mortality (1% CP) compared to our best-estimate values ^27,51^. This change results in a lower baseline severity of SARS-CoV-2 reinfections (6% relative to naïve, Figure 2A). Under this scenario, the severity of first reinfections with novel immune-evading variants are maintained below the severity of naïve infection by pre-existing immunity (Figure 6A). However, the relative change in IFR upon variant emergence is large – up to 5-fold in some scenarios (Figure 6B). As a result, greater immunological protection from severe disease creates vulnerability to sudden changes in nAb potency. These parameter settings have minimal impact on the transmissibility (R_t_) of novel variants at their time of emergence (Figure 6C), with immune evasion conferring rapid initial spread.

**Figure 6:**
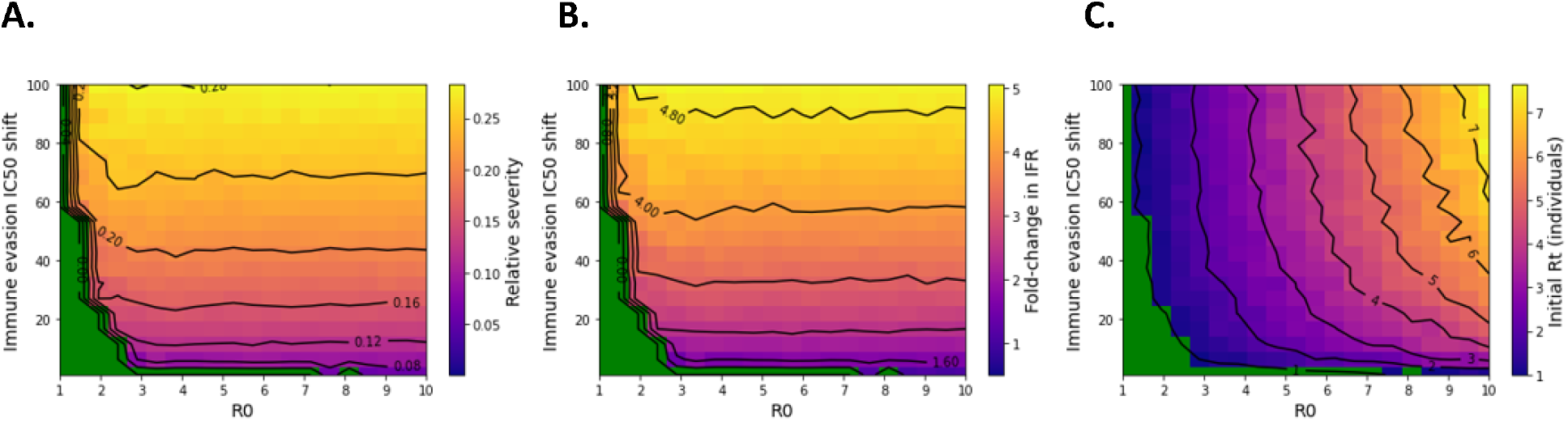
Greater protection from severe disease under endemic spread leads to increased risk of sudden increases in IFR. In these simulations, we used alternative values for endemic model parameters resulting in a lower baseline relative severity of reinfections while varying the R_0_ and immune evasion IC_50_ shift of the invading variant. Green region represents extinction of the invading variant under conditions of pre-existing immunity. **A**. Severity of immune evasive variant infections compared to naïve during the first wave of spread. **B**. Transient fold-change in IFR during immune-evasive variant wave. **C**. R_t_ of the novel variant at the time of introduction.

## Discussion

The work presented here explores the impact of both gradual change and sudden jumps in viral immune evasion upon the population-level immune protection against severe outcomes as a result of COVID-19 infection. We found that, in the absence of immune evasion, natural and vaccinal immunity provide strong protection against severe COVID-19 outcomes. However, with moderate and gradual levels of immune evasion under conditions of antigenic drift, this protection is quickly undermined by a greater number of total infections. Increasing the rate of immune evasion leads to a robust increase in death tolls due to increased infections, rather than an increase in the IFR. A modest rate of immune evasion (t_1/2_= 300 days) is sufficient to double the yearly deaths due to COVID-19. Larger jumps in immune evasion (on the order of 20-40 fold) in new variants due to antigenic shift could lead to large and sudden mortality events (up to a million dead within the United States in a span of 90 days). This would occur due to a transient return of the IFR toward immune-naive levels and a spike in transmission.

The findings presented here are broadly consistent with previously published work by us ^83^, where we showed that even small changes in the IFR under endemic conditions could lead to large death tolls. Here, we have extended that work by demonstrating behavior under excursions from endemicity. We have examined a specific mechanism (antigenic shift leading to large evolutionary decreases in nAb binding potency) and found it leads to increased mortality via a transient change in the IFR or via a spike in the total number of cases. Of course, this is by no means the only mechanism by which SARS-CoV-2 may access increased IFRs. We elucidate the pathways to increased virulence for the virus in a separate literature review ^84^.

Our work has a number of limitations. For simplicity, we did not simulate ongoing inter-strain competitive dynamics. Instead, we assumed that the novel strain emerges into the immunological background induced by the prior strain at steady-state (endemic conditions). We have explored interstrain competition elsewhere ^85^. We also did not model voluntary changes in contact behavior, which is a difficult feature for all epidemiological models to incorporate. Additionally, the sABM model assumes the population is well-mixed and thus does not account for network effects – that is, effects arising from heterogeneity in likelihood of contact between potential interaction pairs. To reduce the impact of these limitations on the results, we focused our analysis on early outbreak dynamics: the R_t_ upon variant emergence.

Our work has exposed the inherent fragility of the current situation and draws out several crucial points for public-health strategy. First, there is a dire need for improved surveillance of viral evolution. Predictive tools linking viral sequence to immune evasion potential are critical at this point, and a number of groups have demonstrated that such approaches are indeed technically feasible ^86–92^. These methods should be deployed at scale-global monitoring of viral evolution and accurate threat forecasting are critical for preventing a sudden mortality event. The IFR of a newly emerged variant is poorly characterized at the time that it emerges, so if a high-IFR variant were to appear, the risk would only become apparent after a large portion of the population was already infected. The time to react to a new variant is not when hospitals and morgues are running out of capacity.

Second, we should seek to reduce the rate of viral evolution by slowing the rate of spread. The current public health position has framed a false dichotomy in the public’s mind about accepting untrammeled viral spread or being subjected to lockdowns. In a previous work, we have shown that a vaccine-only strategy is unable to slow the pace of viral evolution ^65^. At this point, we understand the epidemiology of SARS-CoV-2 to implement multilayered NPIs that can reduce spread, thereby supporting the vaccines in mitigating the death and disability burden of COVID-19. Improvements in indoor air quality, reinstatement of testing-and-tracing protocols, masking requirements in public spaces are all potential options in a toolbox of interventions to achieve this end ^93–95^.

Third, better biomedical interventions, particularly interventions that prevent infection and/or are robust to viral evolution, are urgently needed at this point. For example, nasally-administered antivirals for prophylaxis and early treatment could support the NPIs above in reducing spread. Several such antivirals are currently available in some countries, and have some clinical data supporting their use ^96–100^. Scaling up the production of these treatments and committing to research for additional such antivirals is a low-risk measure that can yield benefits in the short term. On the therapeutic front, with the loss of the monoclonal antibodies ^101,102^, the toolbox for limiting mortality at present is limited to a handful of drugs ^103^.

Investment in better vaccines is also required-intranasally administered vaccines that can generate some degree of mucosal immunity would be beneficial ^104,105^. Better outcomes may also be achievable with the existing vaccines-in a recent work, we point out that repeat dosing with boosters may restore both VEs and VEi ^27^. While several caveats apply to that finding, it speaks to the need for investing in a quantitative understanding of the dynamics and breadth of the nAb response and the risk of tolerability arising from more frequent boosting of the existing vaccines.

The outset of the COVID-19 pandemic was marked by widespread concerns about “doomsday predictions’’ and “fearmongering” ^106–109^. Nevertheless, the outcomes that societies have collectively accepted with the COVID-19 pandemic today exceed the most pessimistic scenarios predicted in the spring of 2020, in terms of death tolls ^110,111^, and duration of impact ^112–118^. With that said, our current reality was predictable, if one started from accurate assumptions. We used model-based approaches to predict the rapid pace of evolutionary immune evasion ^55^, the inability of vaccines to enable a return to pre-pandemic conditions ^119^, the tendency of noncompliance with mitigation measures to incentivize further noncompliance ^120^, and the rapid variant-driven rebound observed upon premature relaxation of mitigation measures ^121^. In each of these cases, our predictions were made many months in advance of the events ^122–125^, and we were by no means the only group to make accurate predictions on these topics ^126–129^.

At its heart, the problem is one of risk management-plausible risks do not need to be inevitable in order to warrant mitigation. In his book “The Black Swan” ^130^, a classic in the risk-management community, author Nassim Nicholas Talib describes a type of event known as a Grey Swan - a rare and highly consequential event that, unlike absolutely unforeseen Black Swan events, can be expected. Our work here shows that ahistorical and potentially destabilizing mortality events as a result of the COVID-19 pandemic are Grey Swans, very much within the realm of the possible. In this paper, we describe one mechanism by which this can occur-antigenic shift (a sudden jump in immune evasion) leading to a reversion to a higher death rate. Such an event, if it were to happen, would not only have been predictable, but can occur repeatedly in the absence of further corrective measures.

While it is often said that “learning to live with the virus” is inevitable, our work suggests that that choice frames a fragile bargain, a deal that can be voided by the virus with little to no prior notice. The levers at our collective disposal can substantially reduce the risk of such an event before it happens.

## Methods

In this analysis, we reimplemented a simplified agent-based model (sABM) developed in our prior work ^27,51^ with modifications to permit simulation of the behavior of invasive immune-evading strains. In the original implementation, the sABM assumes a continuous rate of immune evasion – that is, a continuous rate of nAb potency decay due to antigenic drift. However, viral evolution may proceed in a punctuated fashion, with the emergence of immune-evading variants with significant ability to evade pre-existing immunity ^77,131–133^. To investigate novel immune-evading variants’ propensity to spread and impact on pre-existing protection against severe disease, we simulated the behavior of these variants in the context of pre-existing immunity under endemic SARS-CoV-2 spread.

The original sABM integrates functionality from epidemiological SIRS (susceptible-infected-recovered-susceptible) and nAb pharmacokinetics/ pharmacodynamics (PK/PD) models. The model tracks SARS-CoV-2 reinfections and death risk in a simulated well-mixed population with time-variant nAb titers ^27^. In the epidemiological portion of the model, active infections are tracked. The number of active infections increases by one for each successful exposure and decreases over time at a rate proportional to the recovery period. Exposures occur at a rate proportional to the number of active infections and the intrinsic reproductive number, R_0_. Individuals are randomly drawn from the population for exposure at a probability proportional to their contact rate. Upon exposure, each individual’s probability of infection is determined by their nAb titer according to the published concentration-response (PD) relationship ^134^. Exposure then successfully results in infection on a stochastic basis. Each successful infection decreases the infected individual’s likelihood of survival according to their age-dependent IFR and level of protection from mortality afforded by their nAb titer ^134^. The model also simulates nAb PK: each individual’s antibody titers wane over time according to their nAb half-life and the rate of immune evasion, increase by a fixed multiple upon vaccination if the individual is vaccinated, and increase by a fixed multiple upon successful reinfection.

The original sABM model relies on SIRS epidemiological dynamics. As we sought to develop an understanding of outbreak kinetics and severity upon the emergence of novel immune-evading strains, we re-implemented the model using an SEIRS framework. The latency period (when an individual is developing an infection after exposure, but not yet infectious) may somewhat slow burgeoning outbreaks, reducing the fraction of the population impacted ^135^. The latency period (1/γ) for SARS-CoV-2 was estimated to be 3 days ^136^. Thus, in this implementation of the sABM, individuals who are successfully exposed increase the count of pre-infectious individuals. Pre-infectious individuals become infectious (active infections) at a rate of γ. This model was used to generate Figures 1-3, which explore disease severity under endemic SARS-CoV-2 conditions assuming a continuous rate of immune evasion.

To quantify the impact of novel immune-evading variants, we estimated the R_t_ (effective reproductive number) at the beginning of the outbreak and the relative severity of first-time infections with the novel variant during the outbreak. The R_t_ is the number of secondary infections generated by each index case under current immunological conditions. In the model, the immunological conditions at the time of variant introduction represent endemic conditions for the prior variant. We assumed the prior variant has an R_0_ of 8.2 ^81^. Endemic immunological conditions were reached by running the simulation for 1,000 days. After this point, the novel immune-evading variant was introduced. We assumed 100 individuals are infected with the novel variant at the start of the simulation (100 index cases) out of a total population of 100,000 (0.1% incidence). We calculated the R_t_ over a single simulation day as follows:

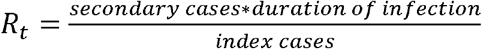

When the duration of infection is 10 days and the secondary cases are the total new infections over the first simulation day after variant introduction. To limit stochastic effects, we replicated the simulation 10 times and calculated the average R_t_.

We also estimated the severity of immune-evasive variant infections relative to naïve infections during the simulated variant-driven spike in transmission. To capture this transient increase in transmission, we ran the simulation for 90 days. We assumed that individuals are immune against reinfection by the novel variant for the duration of the simulation. The relative severity of these infections are determined based on an individual’s antibody titer at the time of infection:

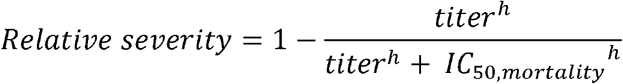

where titer is an individual’s nAb titer, h is the Hill coefficient, and IC_50,mortality_ is the nAb titer required for 50% protection from COVID-19 death. We also report the fold-change in apparent IFR upon emergence of the variant, which is the ratio of the immune-evasive outbreak relative severity to the relative severity under endemic conditions.

IC_50,mortality_ and and IC_50,infection_ are parameterized based on nAb concentration-COVID-19 protection relationships published in Khoury et al ^134^. We note that in the published model, protection levels from mild symptomatic infection and severe COVID-19 are evaluated. We assume that the nAb titer required for protection from mild symptomatic infection (IC_50,mild_) is lower than that required to provide sterilizing immunity (IC_50,infection_). As a result, the precise value for IC_50,infection_ is not known, but IC_50,mild_ is the lower bound. Additionally, the peak convalescent nAb titer (1 CP) is known to confer at least 80% protection from reinfection ^137^, implying that IC_50,infection_ must fall below this titer. Thus, the plausible range for IC_50,infection_ is 0.2 - 1 CP. Similarly, the Khoury study estimates the nAb titer required for 50% protection from severe disease (IC_50,severe_), which includes hospitalizations, ICU cases, and deaths ^134^. As death is the most severe COVID-19 outcome, we assumed that IC_50,severe_ is an upper bound for IC_50,mortality_. We explored scenarios in which IC_50,mortality_ is up to 10-fold lower than IC_50,severe_.

## Data Availability

All data produced in the present study are available upon reasonable request to the authors

## Notes

### Competing Interest Statement

MS, LY, and AC are employees of Fractal Therapeutics, Inc.

### Funding Statement

This study did not receive any funding

### Author Declarations

The study used ONLY openly available human data that were originally located at https://academic.oup.com/cid/article/73/3/e531/5880016?login=false

### Summary of Updates

This version of the manuscript has been revised to update the following: Original DOI link for this manuscript at https://doi.org/10.1101/2023.02.26.23286471 is broken

