## Supplementary Materials for "The gray swan: model-based assessment of the risk of sudden failure of hybrid immunity to SARS-CoV-2"

**Affiliations:**

**Abstract:**

In the fourth year of the COVID-19 pandemic, public health authorities worldwide have adopted a strategy of learning to live with SARS-CoV-2. This has involved the removal of measures for limiting viral spread, resulting in a large burden of recurrent SARS-CoV-2 infections. Crucial for managing this burden is the concept of the so-called wall of hybrid immunity, through repeated reinfections and vaccine boosters, to reduce the risk of severe disease and death. Protection against both infection and severe disease is provided by the induction of neutralizing antibodies (nAbs) against SARS-CoV-2. However, pharmacokinetic (PK) waning and rapid viral evolution both degrade nAb binding titers. The recent emergence of variants with strongly immune evasive potential against both the vaccinal and natural immune responses raises the question of whether the wall of population-level immunity can be maintained in the face of large jumps in nAb binding potency. Here we use an agent-based simulation to address this question. Our findings suggest large jumps in viral evolution may cause failure of population immunity resulting in sudden increases in mortality. As a rise in mortality will only become apparent in the weeks following a wave of disease, reactive public health strategies will not be able to provide meaningful risk mitigation. Learning to live with the virus could thus lead to large death tolls with very little warning. Our work points to the importance of proactive management strategies for the ongoing pandemic, and to the need for multifactorial approaches to COVID-19 disease control.

**Supplementary Figures**

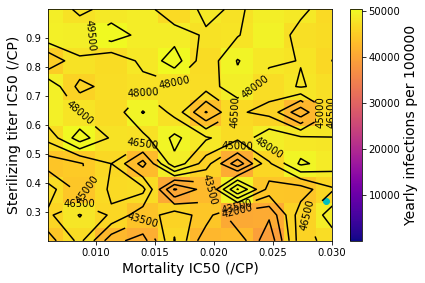

**Figure S1:** Yearly SARS-CoV-2 infections under conditions of endemic spread in the absence of immune evasion. Blue point represents best estimate parameter values.

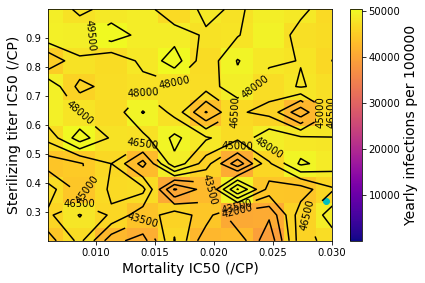

**Figure S2:** Yearly SARS-CoV-2 infections under conditions of endemic spread with immune evasion. Blue point represents best estimate parameter values.
**A. B. C.**

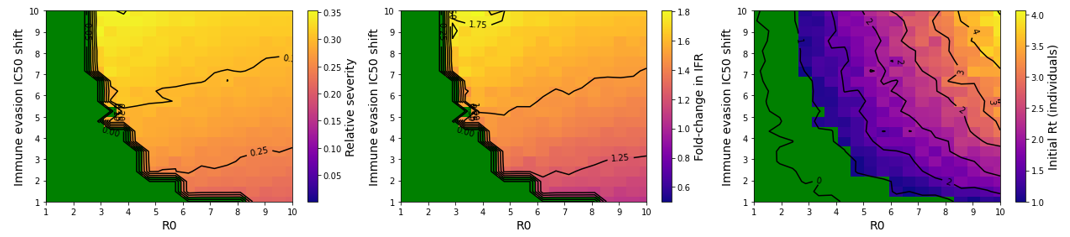

**Figure S3:** Immune evading variants with smaller nAb potency drops may spread efficiently with minimal impact on apparent IFR. This figure is a cropped version of Figure 4, with the y-axis covering nAb potency shifts of 1 to 10-fold. Green region represents extinction of the invading variant under conditions of pre-existing immunity. **A.** Severity of immune evasive variant infections compared to naïve during the first wave of spread. **B.** Transient fold-change in IFR during immune-evasive variant wave. **C.** R_t_ of the novel variant at the time of introduction.

**A. B. C.**

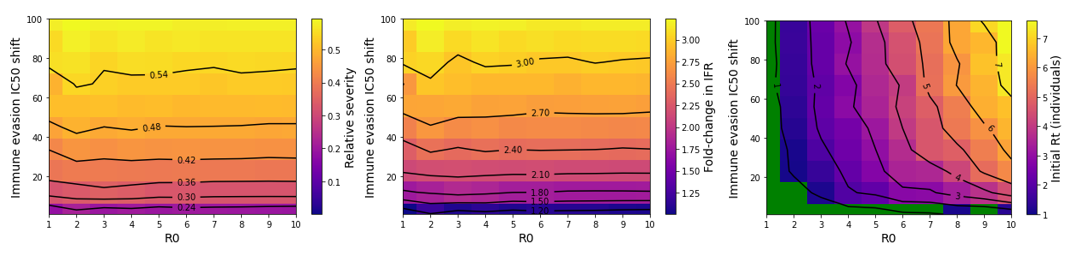

**Figure S4:** Outcomes of immune evasive variant invasion in the absence of vaccines. This figure mirrors Figure 4, with no vaccines. Green region represents extinction of the invading variant under conditions of pre-existing immunity. **A.** Severity of immune evasive variant infections compared to naïve during the first wave of spread. **B.** Transient fold-change in IFR during immune-evasive variant wave. **C.** R_t_ of the novel variant at the time of introduction.

**A. B. C.**

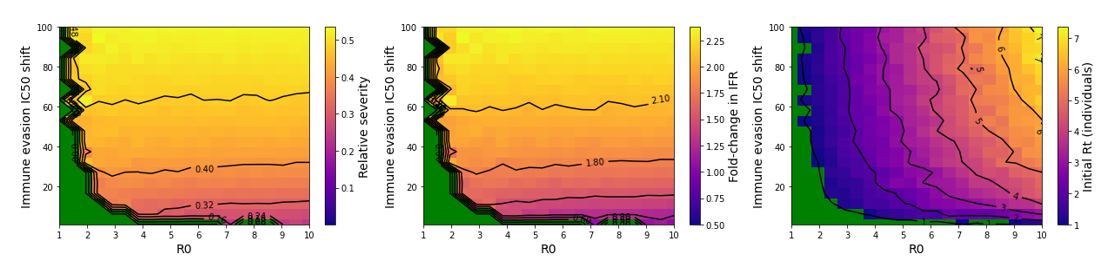

**Figure S5:** Increased vaccination mitigates some of the impact of immune evasive variants on mortality. This figure mirrors Figure 4, with an improvement in vaccination conditions to 90% compliance and twice-yearly boosting. Green region represents extinction of the invading variant under conditions of pre-existing immunity. **A.** Severity of immune evasive variant infections compared to naïve during the first wave of spread. **B.** Transient fold-change in IFR during immune-evasive variant wave. **C.** R_t_ of the novel variant at the time of introduction.
